# Respiratory Exacerbations Increase with Chronic PM_2.5_ Exposure in Current and Former Smokers

**DOI:** 10.1101/2025.05.27.25328449

**Authors:** James L. Crooks, Zhongying Wang, Morteza Karimzadeh, Surya P. Bhatt, Craig P. Hersh, Dawn L. DeMeo, David Baraghoshi, Min Hyung Ryu, Elizabeth A. Regan

## Abstract

**Rationale:** Short-term exposure to fine particulates (PM_2.5_) transiently increases the risk of respiratory exacerbations, but the contribution of chronic, long-term particulate exposure to respiratory exacerbations is poorly defined.

**Objectives:** To assess long-term effects of PM_2.5_ exposure on risk of severe respiratory exacerbations.

**Methods:** A longitudinal cohort of current and former smokers with and without COPD were surveyed every six months for severe exacerbation events. PM_2.5_ concentrations at participant addresses were estimated using satellite, reanalysis, and ground-based monitoring data sources.

**Measurements and Main Results:** The relative risk of severe exacerbation increased by a factor of 1.516 (CI: 1.226, 1.873; *p* = 0.00012) for every 10 μg/m^3^ increase in long-term PM_2.5_ exposure across all participants. The effect in the non-COPD participants was greater, with a relative risk of 2.639 (CI: 1.840, 3.756; p<0.0001). Significant effect modifiers with greater effect of PM_2.5_, included prior exacerbations, female sex, and neighborhood characteristics and as well as smoking status, white race, disease severity, asthma diagnosis, and age at enrollment. Significant positive associations for PM_2.5_ on exacerbations were identified at levels below the EPA primary annual standard for PM_2.5_ of 9.0 μg/m^3^.

**Conclusions:** Persistent exposure to fine particulates is a significant risk factor for severe respiratory exacerbations in current and former smokers, and in patients with or at risk of COPD. The effect of fine particulates on the risk of severe exacerbations appears to be greater in those current and former smokers without COPD. The EPA annual PM_2.5_ standard may be inadequate to prevent ongoing lung injury.

## Background

Heavy cigarette smoking is a well-known risk factor for Chronic Obstructive Pulmonary Disease (COPD) ^1^, and is associated with extensive burden of disease and increased healthcare costs ^2^. However, other inhaled exposures are also commonly associated with both respiratory illness and COPD, including biomass exposure, occupational exposures, and importantly air pollution. Both current and former smokers, as well as never smokers, experience long-term health impacts from air pollution^3^. Ozone, NO_2_, and particulate matter with an aerodynamic diameter less than 2.5 microns (PM^2.5^), have adverse effects on health including increased mortality ^4,5^, cardiovascular disease^6^, progression of emphysema and reduced lung function^7, 8,9^. Air pollution, especially its gaseous components, have been linked to risk of COPD incidence, prevalence and progression ^1,10,11^. More recent work has defined the importance of small particulates (PM_2.5_) in air pollution that associate with reduced lung function and COPD ^11^. An important factor for understanding a potential role for air pollution and COPD disease progression are episodes of respiratory exacerbations from various causes that are associated with disease progression in COPD^12,13^. Severe exacerbations are defined as those that require hospitalization, emergency room visits, and antibiotic and/or steroid treatment. Respiratory exacerbations also impact never-smokers and smokers without COPD, particularly with short term changes in air quality^14,15^. Moreover, the impacts of combined pollutants – ozone, particulates and NO_2_ are well documented^16–18^, However, the singular role of PM_2.5_ exposure on the risk of acute respiratory exacerbations is less well defined, especially in smokers with a diagnosis of COPD. Further, long- vs short-term exposures to particulates are suggested to have different risks, with evidence of recovery following short-term exposures ^19^.

Air pollution intensity and constituents vary by location; and an individual’s residential location along with severity of local air pollution may be an important social determinant of health related to COPD^20,21^. Determining the sources and impact of various factors on disease risks is critical to improving outcomes of care for COPD and other respiratory diseases. Smoking cessation, for example, is strongly recommended for improved health. Still, persistent exposure to outdoor air pollution may remain as a precipitating factor for exacerbations of disease in both current and former smokers. Other, potentially modifiable factors that affect the risk of exacerbations associated with air pollution are important to define and require detailed individual information as well as the timing and duration of exposures and the impact of specific pollution components. We studied the impact of chronic, local PM_2.5_ concentrations on respiratory exacerbations in a well-characterized established longitudinal cohort of current and former smokers with and without COPD -the COPDGene study, over a period of 16 years (2006-2021). We hypothesized that increased long-term exposure to fine particulate matter would be associated with increased risk of exacerbations. This work was approved by the National Jewish Health Institutional Review Board (HS-3752).

## Methods

### Population

The COPDGene Study enrolled 10,652 individuals at 21 clinical centers around the US who had at least a 10-pack-year history of smoking and the details have been published^22^. The COPDGene research protocol was approved by the institutional review board at each of the 21 clinical centers, and all participants provided written informed consent. Current residential address was collected from a limited number of the 10,305 participants enrolled in Phase 1 of the study (2007-2012) and the majority of participants in Phase 2. (2012-2017). A small group of never smokers (n=454) were enrolled in Phase 1 and Phase 2 as a control group, and current address was collected from all remaining participants. Thus, a patient’s enrollment may have occurred either in Phase 1 or Phase 2. In Phase 3 (2017-2023), updated address information was collected from participants. Jittered patient locations for all phases are visualized in Figure XXX, colored by their enrollment clinical center.

**Figure 1.**
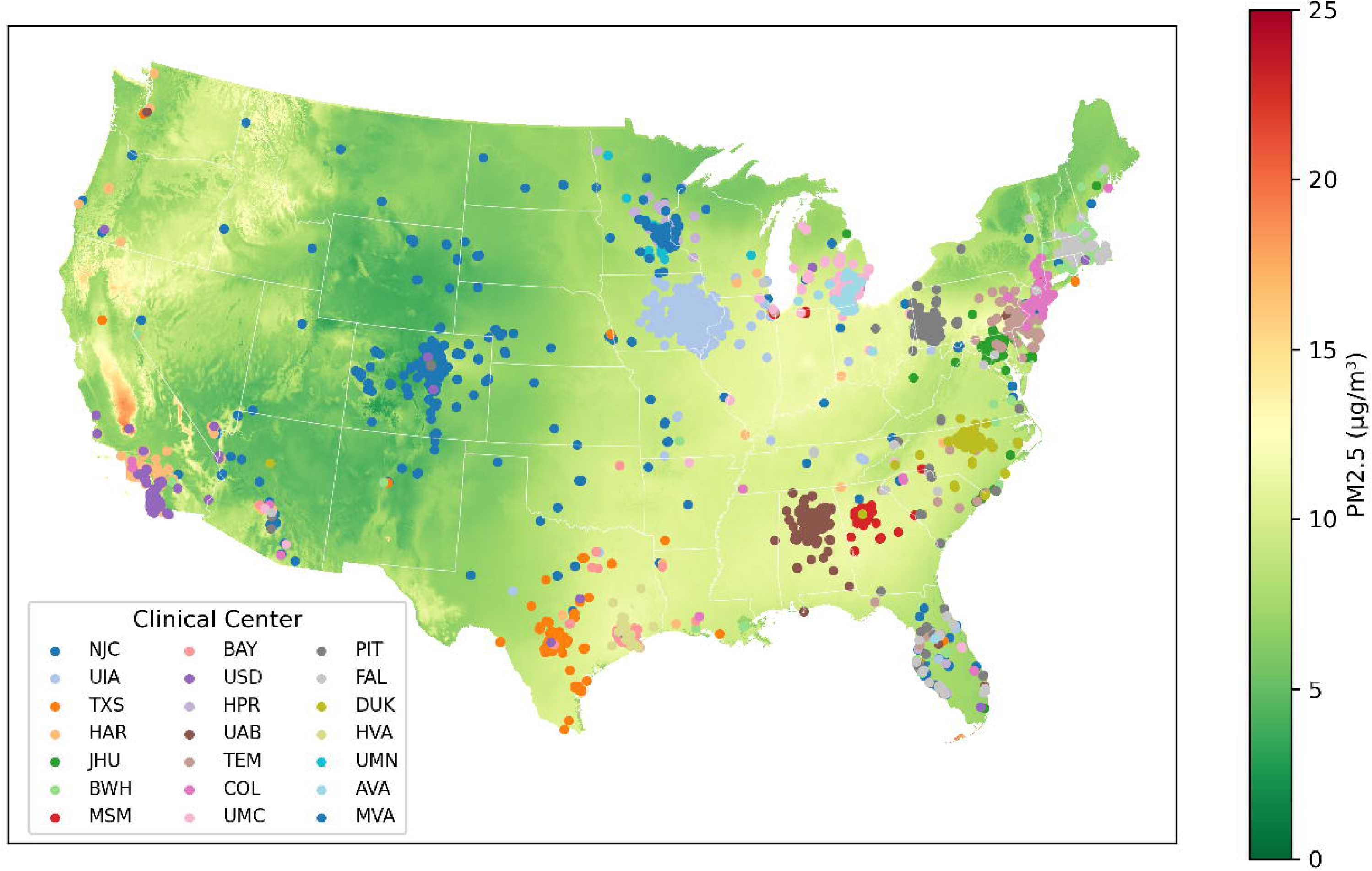
Population Locations and PM2.5 Concentrations. Map of geocoded participant residence locations for all phases overlaid on 16-year average PM2.5 concentrations from the GeoAI model. Residence locations have been jittered to protect privacy. Participant locations are color coded by the COPDGene cinical site at enrollment, though in some cases participants move between phases.

### Exacerbations

Participants in COPDGene were surveyed approximately every six months by phone or email, and asked about respiratory exacerbations that resulted in an emergency room visit or hospitalization and was accompanied by treatment with antibiotics and/or glucocorticoids and occurred “since we last spoke”. Survey collection started after Phase 1 was initiated, with the first surveys administered in May 2009. A total of 103,017 of these longitudinal follow-ups (LFU) phone surveys were collected from 9,479 participants during our study period ending in December 2021. A total of 6,510 participants had at least one geocoded address and at least one phone survey; this subset of participants had 86,692 unique surveys. Of the 10,652 participants in the COPDGene cohort, a total of 1,573 addresses were obtained in Phase 1, 6,298 addresses in Phase 2, and 4,455 addresses in Phase 3, encompassing 6,719 unique individuals, and geocoded using ArcGIS Pro 2.8.0^23^.

### Particulate Matter

Daily PM_2.5_ concentrations were previously estimated at a 1km spatial resolution raster over the contiguous U.S. for the years 2005-2021 using a geospatial artificial intelligence (GeoAI) model ^24^. These daily concentrations were estimated back approximately eighteen months prior to the beginning of Phase 1 to allow calculation of long-term exposures. The GeoAI model incorporated PM_2.5_ monitor observation, satellite aerosol optical depth (imputed where missing), global reanalysis products, weather data, and elevation. Figure XXX shows the spatial distribution of the 16-year average of the model’s daily PM_2.5_ predictions.

A full 16-year time-series of daily PM_2.5_ concentrations was assigned to each participant’s geocoded residences using the raster cell into which the residency fell. These concentrations were then averaged over the time intervals between each consecutive pair of surveys. However, following a protocol change on January 1, 2014, whenever a study visit occurred between surveys the average was computed over the interval from the date of the study visit to the date of the survey rather than the interval between the surveys.

Details of how these issues were handled are provided in the supplemental methods. A total of 5,638 participants with 41,735 surveys had at least one assignable PM_2.5_ exposure and were used in the analyses below. Because all time intervals were at least 59 days long, the average PM_2.5_ concentrations are all considered ‘long-term exposures’ as defined by the U.S. Environmental Protection Agency^25^

### Neighborhood Characteristics

Geocoded locations were used to identify US census tracts in which participants lived and thereby derive their tracts’ 2018 area deprivation index (ADI)^26^, 2018 social vulnerability index (SVI)^27^, and neighborhood’s Homeowners Loan Corporation (HOLC) grade from the 1930s (i.e., “redlining” grade)^28^.

### Statistical Analyses

Following geocoding, all subsequent analyses were performed in R version 4.4.2^29^.

### Crude Model

Associations between the ambient long-term average PM_2.5_ concentrations and self-reported exacerbation counts during the between-survey intervals were estimated using quasi-Poisson models implemented in the gnm^30^ package, with a separate fixed stratum intercept for each subject and an offset term accounting for the logarithm of the between-survey interval length. The fixed intercept accounts for all individual-level factors at enrollment including prior medical history (e.g., asthma diagnosis, prior exacerbations) as well as time-invariant factors (e.g., race). A crude model included only strata intercepts and PM_2.5_ as predictors.

### Primary Model

Our primary model included these plus potential confounders. Because COPD can be characterized by low blood oxygen levels, the primary model controlled for residence elevation. Likewise, since COPD tends to be a progressive disease, the primary model also included nonlinear control for time using linear and quadratic time terms. Model results with p<0.05 were considered ‘statistically significant’.

### Effect Modification Analyses

To determine whether associations identified in our main model might be impacted by meteorology, time period, neighborhood and demographic factors, and medical history at baseline, we explored the modification of the PM_2.5_ effect. Details are provided in the Supplementary methods.

Furthermore, since the U.S. EPA annual standard for PM_2.5_ is currently 9.0 μg/m^3^, to evaluate whether the standard is sufficiently protective, the analysis was repeated using only surveys associated with addresses where the 16-year and between-survey average PM_2.5_ concentrations were below 9.0, 8.5, 8.0, 7.5, and 7.0 μg/m^3^.

## Results

### Participants

Demographic characteristics of the cohort at enrollment are given in Table 1. The study included 5,638 participants, of whom 2,756 were male and 2,882 female. The racial distribution included 4,123 whites and 1,515 Black/African-Americans. The mean age was 60 years. Most participants had some college education or were college graduates. On average, participants had a BMI of 28.2, an FEV_1_/height^2^ of 0.83 L/m^2^, an FEV_1_ and FVC of 2.34 and 3.29 liters, respectively, an FEV_1_/FVC of 0.73, a 6-minute walk distance of 1,476 feet, a SGRQ total score of 15, percent emphysema of 2% (% LAA@-950), and a median MMRC dyspnea score of 1.05 with 54% of the participants reporting a score of zero. Smoking characteristics at enrollment found that a small majority were former smokers (50%), with current smokers 43% and never smokers 7%, and 92% had never experienced a severe respiratory exacerbation. In addition, nearly half (44%) were classified as not diagnosed with COPD based on spirometry, which we have termed “GOLD 0”. Fewer than half of participants reported a co-morbid disease, with a history of asthma reported in 17%, congestive heart failure 2% and coronary artery disease 6%.

**Table 1.**
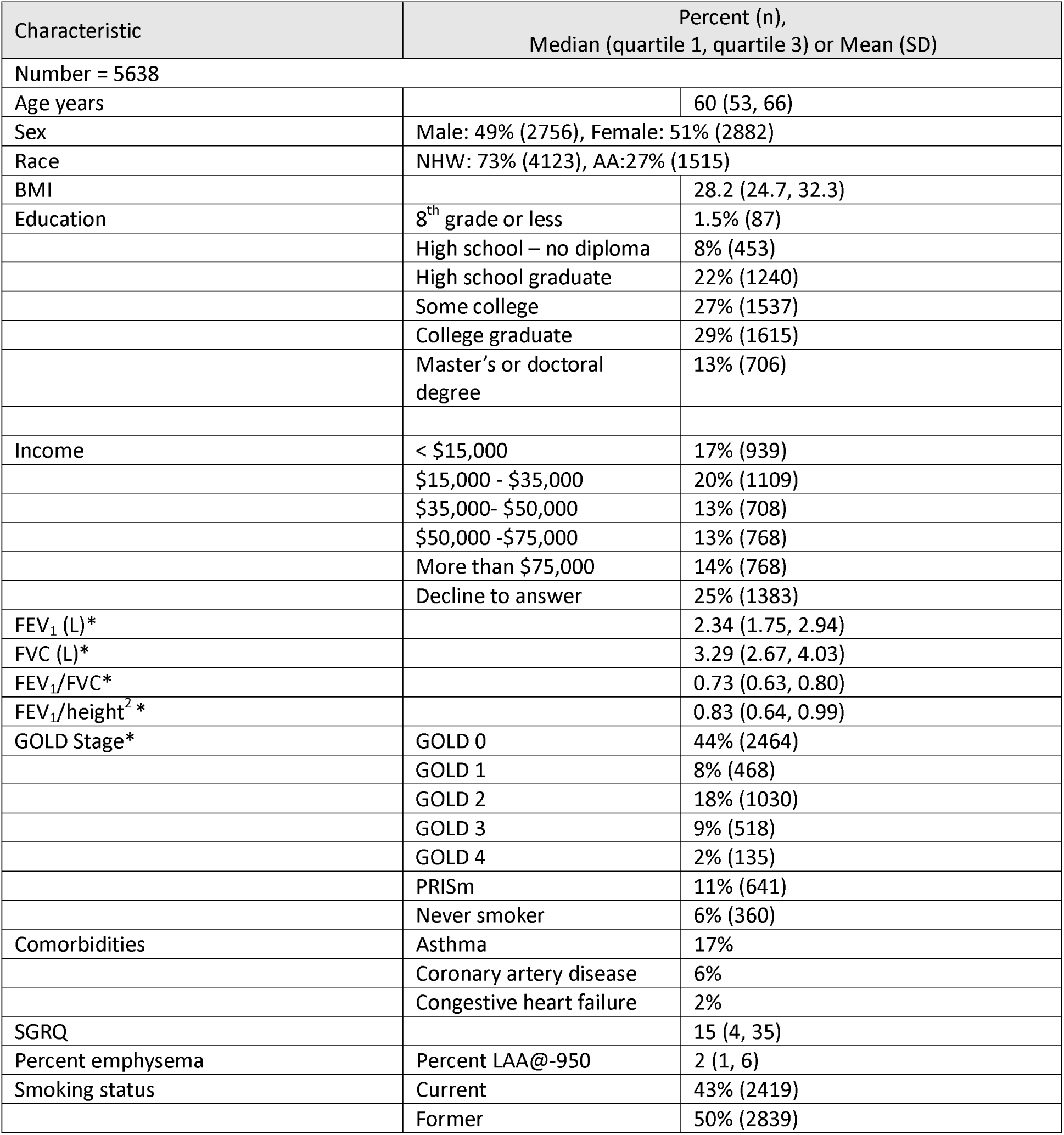
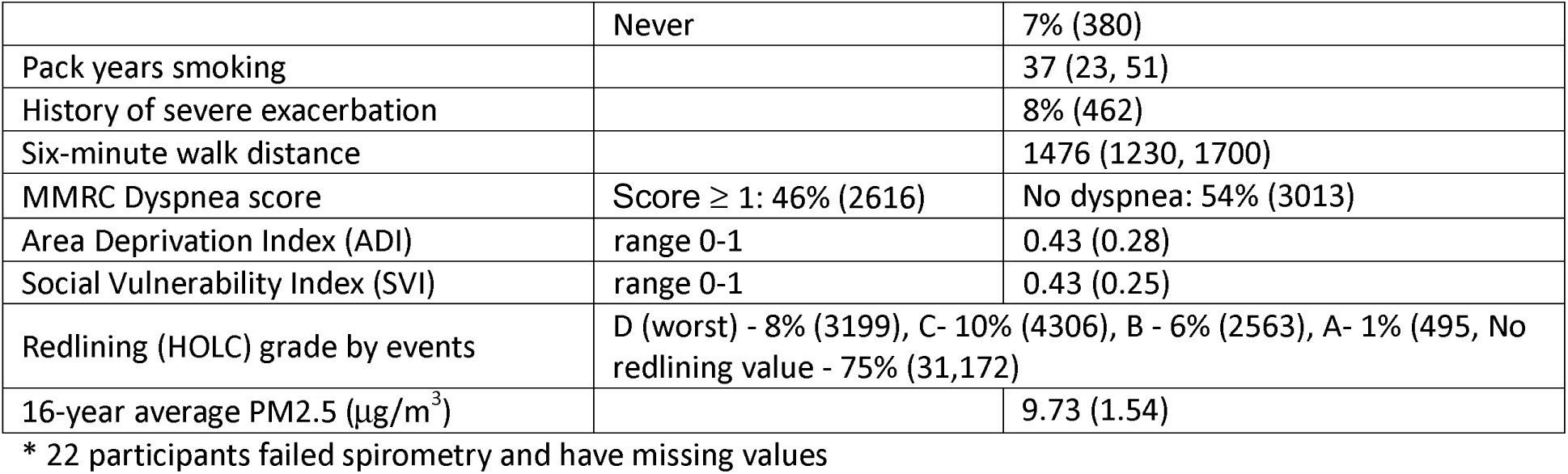
Population Characteristics.

The average number of reported between-survey exacerbations was 0.20 per participant, while the average between-survey interval duration was 205 days. Exacerbation rates were heavily skewed, with median of 0/year, a mean of 0.359/year, and a maximum of 12/year (Supplemental Figure S2). Fully 3630 (64.4%) participants had no exacerbations. The time intervals between surveys were likewise skewed, with a median of 0.556 years (203 days), a mean of 0.647 years (237 days), and a maximum of 10.27 years (3752 days) (Supplemental Figure S2). The average PM_2.5_ concentration between surveys was 8.81 μg/m^3^, and the average 16-year PM_2.5_ across surveys was 9.73 μg/m^3^. Average ADI and SVI across surveys were both 0.43 on a scale of 0-1. While most geocoded addresses did not fall into a neighborhood with a “redlining” or Homeowners Loan Corporation (HOLC) grade, of those that did, most were in the lowest two grades (C and D). Looking at population characteristics by reported exacerbations, 3630 participants reported no episodes of severe exacerbation, 1337 reported up to one per year and 671 reported more than 1 per year (Table 2). Those who reported any severe exacerbation had higher (worse) ADI and SVI

**Table 2.** Characteristics and Exposures by Exacerbation Rates.

| Characteristic | Exacerbations |  |  |
| --- | --- | --- | --- |
|  | None | 0-1 per year | > 1 per year |
| Average PM 2.5 exposure ($\mu\text{g}/\text{m}^3$) | 8.85 (1.52) | 8.80 (1.39) | 8.95 (1.64) |
| Age | 60 (9) | 61 (8) | 59 (9) |
| Sex (% male) | 52% | 42% | 45% |
| Race (% NHW) | 72% | 78% | 69% |
| Smoking status |  |  |  |
| Current | 43% | 41% | 47% |
| Former | 48% | 56% | 53% |
| Never | 9% | 3% | 2% |
| ADI | 0.44 (0.25) | 0.44 (0.25) | 0.47 (0.25) |
| SVI | 0.47 (0.28) | 0.46 (0.28) | 0.54 (0.28) |

### Crude and Primary Models

Our crude model estimated a factor of 1.553 (95% CI: 1.293, 1.865; p <0.0001) increase in exacerbation risk per 10μg/m^3^ increase in between-survey average PM_2.5_ concentration. Our primary model estimated a factor of 1.516 (CI: 1.226, 1.873; *p* = 0.00012) increase for the same change in concentration.

### Effect Modification Analyses

Table 3 presents our most important effect modification results. The inclusion of linear and interaction terms for race, gender, GOLD stage, prior severe exacerbations, SVI, and ADI improved overall model fit based on the likelihood ratio test. Among these, statistically significant differences were observed between Blacks and whites (*p* = 0.0001, higher for Whites), males and females (*p* = 0.0102, higher for females), prior vs. no prior exacerbations (*p* = 0.0016, higher for prior), and between PRISm and GOLD 2 vs. GOLD 0 (*p* = 0.0011 and *p* < 0.0001, respectively, higher for GOLD 0 in both cases). Furthermore, in stratified results, GOLD 0 yielded the strongest association with PM_2.5_ (RR = 2.629; 95% CI: 1.840, 3.756; *p*<0.0001) among all GOLD stages. Within GOLD 0, former smokers had a stronger PM_2.5_ association (RR = 3.245; 95% CI: 2.030, 1.185; *p* < 0.0001) than current smokers (RR = 2.015; 95% CI: 1.185, 3.426; *p* = 0.0097). While smoking status was not statistically significant under the likelihood ratio test, a statistically significant association was observed among non-current smokers (RR = 1.686; 95% CI: 1.298, 2.191; *p* < 0.0001). The PM_2.5_ association decreased with increasing ADI. However, while SVI was significant according to the likelihood ratio test, the interaction term itself was not, indicating that the improvement in model fit was driven by the linear term.

**Table 3.** Effect Modifiers.

| Modifier | Likelihood ratio<br>test p-values | Interactions |  |  | Point values |  |  |
| --- | --- | --- | --- | --- | --- | --- | --- |
|  |  | Comparison | Estimate (95% CI)* | p-value | Category | Estimate<br>(95%CI)* | p-value |
| Race | **0.0001** | **Black vs White** | **0.420 (0.271, 0.652)** | **0.0001** | **White (N=4123)** | **1.838 (1.458, 2.318)** | **<0.0001** |
|  |  |  |  |  | Black (N=1515) | 0.772 (0.516, 1.165) | 0.21 |
| **Sex** | **0.0102** | **Female vs Male** | **1.657 (1.127, 2.435)** | **0.0103** | Male (N=2756) | 1.103 (0.799, 1.524) | 0.55 |
|  |  |  |  |  | **Female (N=2882)** | **1.828 (1.417, 2.357)** | **<0.0001** |
| Smoking status^ | 0.32 | Former vs Current | 0.756 (0.517, 1.107) | 0.1505 | Current (N=2419) | 1.275 (0.933, 1.743) | 0.1268 |
|  |  | Never vs Current | 1.339 (0.235, 7.643) | 0.7424 | **Former (N=2839)** | **1.686 (1.293, 2.191)** | **<0.0001** |
|  |  |  |  |  | Never (N=380) | 2.258 (0.403, 12.65) | 0.35 |
| GOLD Stage^ | **0.0003** | **PRISm vs GOLD 0** | **0.343 (0.180, 0.652)** | **0.0011** | **GOLD 0 (N=2464)** | **2.629 (1.840, 3.756)** | **<0.0001** |
|  |  | GOLD 1 vs GOLD 0 | 0.503 (0.218, 1.159) | 0.1067 | PRISm (N=641) | 0.901 (0.516, 1.573) | 0.7143 |
|  |  | **GOLD 2 vs GOLD 0** | **0.339 (0.204, 0.562)** | **<0.0001** | GOLD 1 (N=468) | 1.322 (0.614, 2.849) | 0.4743 |
|  |  | GOLD 3 vs GOLD 0 | 0.764 (0.448, 1.305) | 0.3252 | GOLD 2 (N=1030) | 0.890 (0.607, 3.032) | 0.5508 |
|  |  | GOLD 4 vs GOLD 0 | 0.468 (0.199, 1.102) | 0.0824 | **GOLD 3 (N=518)** | **2.010 (1.319, 3.032)** | **0.0012** |
|  |  |  |  |  | GOLD 4 (N=135) | 1.231 (0.560, 2.704) | 0.6054 |
|  |  | Never smokers vs GOLD 0 | 0.898 (0.141, 5.734) | 0.9098 | Never smokers (N= 360) | 2.362 (0.388, 14.58) | 0.3546 |
| **Prior severe exacerbation^** | **0.0016** | **Yes vs No** | **2.085 (1.324,3.286)** | **0.0015** | **No (N= 5176)** | **1.314 (1.044, 1.655)** | **0.0199** |
|  |  |  |  |  | **Yes (N=462)** | **2.74 (1.798, 4.180)** | **<0.0001** |
| **SVI** | **0.0094** |  | 0.759 (0.386,1.494) | 0.4246 |  |  |  |
| **ADI** | **0.0052** |  | **0.312 (0.152, 0.639)** | **0.0014** |  |  |  |

\* Per 10 $\mu\text{g}/\text{m}^3$ increase in $\text{PM}_{2.5}$
^ evaluated at enrollment

Supplemental Table S2 presents further effect modification results. Models with interactions by 16-year average PM_2.5_ (*p* = 0.0003) and age at enrollment (*p* < 0.0001) yielded a significant improvement in model fit based on the likelihood ratio test. Among those, age had a statistically significant interaction coefficient. However, the PM_2.5_ association strengthened with age (Supplemental Figure 7), implying that its improvement in fit was driven by the linear term.

For each continuous modifier, plots of the main PM_2.5_ association over the range of that modifier are given in Supplemental Figures S4-S13. These figures show that the association between PM_2.5_ and exacerbations was positive and significant when 16-year average PM_2.5_ fell in the range 2.5–12.5 μg/m^3^ (S4), 16-year average daily minimum temperature in the range 3– 23 °C (S5), residence elevations below 900 meters (S6), SVI below 0.85 (S7), ADI below 0.55 (S8), age at enrollment above 55 years (S9), FEV_1_/height^2^ above 0.5 L/m^2^ (S10), % emphysema below 27% (S11), pack years below 75 (S12), and BMI between 20 and 50 (S13).

Among categorical modifiers (Supplemental Table S2), statistically significant differences were observed between 2014 or earlier vs. 2015-2018 (*p* = 0.0089, higher for 2015-2018) and HOLC grade A vs. D (*p* = 0.0027, higher for grade D). These results indicate that the exacerbation risk associated with PM_2.5_ was higher for participants who were white and female, those with prior exacerbations and less smoking history and disease progression at enrollment, those who were older at enrollment, and those living in neighborhoods with less deprivation now but with more historical disinvestment.

### Sensitivity Analyses

Supplemental Table S3 presents sensitivity results. Associations remained positive under all sensitivity analyses except the model with linear control for time, with risk ratios for the other analyses ranging from 1.263 to 1.561 per 10μg/m^3^ increase in PM_2.5_. The binomial model analyzing presence or absence of exacerbations yielded an odds ratio of 2.097 (95% CI: 1.542, 2.851; *p* < 0.0001), while the mixed effects model yielded a risk ratio of 1.463 (95% CI: 1.167, 1.832; *p* = 0.0009) for the same exposure change. Using only surveys with short interval periods had no qualitative impact on the result (Supplemental Figure S14). Using only surveys taken while subjects lived at residences with 16-year average PM_2.5_ below the current standard yielded a statistically significant positive associations below 9 μg/m^3^, 8.5 μg/m^3^, and 8.0 μg/m^3^ (Supplemental Figure S15). Using only surveys with between-survey average PM_2.5_ below the current standard yielded a statistically a significant positive association below 9 μg/m^3^ (Supplemental Figure S16).

## Discussion

We estimated a positive association between PM_2.5_ exposure and severe exacerbation risk, corresponding to a 51.6% increase in risk for each 10 μg/m^3^ increase in PM_2.5_. In addition, we found that the exacerbation risk driven by PM_2.5_ exposure tended to be higher among populations considered less sensitive or vulnerable in some ways (whites, patients living in less deprived areas, and patients with little disease progression) and more sensitive or vulnerable in other ways (females, older patients, patients with prior exacerbation history, patients living in historically disinvested areas, patients with moderate disease progression). Participants without spirometric obstruction (GOLD 0) at enrollment had the strongest PM_2.5_ association among all subgroups, and, within that group, the former smokers had a stronger association than current smokers. No subgroup of participants had a protective association with PM_2.5_ except the small number (N = 84) living in HOLC grade A neighborhoods. We found little evidence that PM_2.5_ mediates the impact of prior exacerbation history on future exacerbations.

Our mixed results in terms of vulnerability or susceptibility factors belie the complexity of real-world multi-domain exposures through the life course and their potentially countervailing impacts on COPD patients’ sensitivity to air pollution. This work is the first to study the impact of PM_2.5_ on respiratory exacerbations in a well-characterized COPD cohort of this size and opens the door to future studies investigating PM_2.5_ associations with lung imaging measures and molecular endpoints, as well as multi-pollutant impacts.

This work has several limitations. First, because addresses were collected on different days than the phone surveys, we necessarily made an assumption regarding when participants moved between residences in order to assign exposures to between-survey intervals. We explored a different assumption in a sensitivity analysis, but other assumptions are possible. Second, exposures were estimated not measured, so errors in our PM_2.5_ estimates may affect our results; however, the error involved is of the Classical type, which usually yields associations that are less strong than the true association. Furthermore, modeling work on other outcomes has shown that using PM_2.5_ estimated with error yields results similar to the truth^31^.

Beyond potential clinical importance, our results have regulatory implications. A significant positive association was observed in areas with 16-year average PM_2.5_ concentrations below 8.0 μg/m^3^, which is below the current U.S. EPA PM_2.5_ level annual standard, and a significant positive association was also observed when using only between-survey average concentrations below the standard. This indicates that, at least in the population studied here, the standard may insufficient to protect public health. Moreover, the regulation of PM_2.5_ is largely justified based on its relationships with mortality and cardiovascular health effects, not respiratory health effects, so demonstrating a respiratory health effect due to long-term PM_2.5_ exposure further increases the evidence basis for standard setting.

## Data Availability

PM2.5 data used in the present work available online at https://geohai.org/projects/estimating-air-pollution.html.
De-identified, merged analytic datasets will be made available upon final publication.

