## Supplemental Figures and Tables for "Respiratory Exacerbations Increase with Chronic PM_2.5_ Exposure in Current and Former Smokers"

***Detailed Methods, Tables and Figures for***

**Detailed methods:**

Population:

The COPDGene study enrolled 10,652 individuals at 21 clinical centers around the US who had at least a 10-pack-year history of smoking and the details have been published^17^. They were characterized after enrollment with spirometry to determine COPD disease status and severity of lung disease. In order to accomplish genetic studies, enrollment was limited to non-Hispanic White and Black participants, with enhanced enrollment of African-Americans and people with known COPD. Exclusions were pre-existing interstitial lung disease, bronchiectasis and other chronic lung disease, with the exception of asthma. In-person study visits were conducted In each of three phases of the study (Phase 1 2007-2012, Phase 2 2012- 2017, Phase 3 2017- 2022), and subjects underwent a comprehensive health evaluation including spirometry, current and past health issues, respiratory symptoms, medications, and blood samples. During the COVID pandemic (2020- 2021) visits were conducted virtually until in-person visits were permitted, and updated spirometry and CT scans were performed asynchronously from the questionnaires until in-person visits could be resumed.

Spirometry was performed at each in-person study visit according to American Thoracic Society (ATS) standards ^18^, and inspiratory and expiratory chest CT scans were acquired according to a standardized protocol. Quantitative analysis of the CT scans was performed by Thirona (Njimigen, Netherlands) and defined severity of emphysema (LAA -950) as well as other measures. Participants were identified as having COPD based on a post bronchodilator FEV_1_/FVC ratio of <0.7 and classified for severity of disease using GLI Global prediction equations ^19^. For analysis of spirometry as a continuous variable we used FEV1/ height^2^ ^20^. Dyspnea and quality of life were quantified with the modified Medical Research Council (mMRC) dyspnea score ^21^and the St George’s Respiratory Questionnaire^22^ . Severe exacerbation events, smoking status, history and pack-years were identified by questionnaire at enrollment and subsequent study visits.

Geocoding and phone contacts:

Geocoded addresses collected at study visits do not in general correspond to phone visits dates. Furthermore, participants may have moved between Phase, and the dates of any moves were not recorded. Therefore, assumptions regarding when people moved were necessary in order to calculate the average PM_2.5_ concentration over each participant’s between-survey intervals. The method used to do this is shown in Supplemental Table 1. For example, a participant with a Phase 3 geocoded address and an interval starting up to 2 years after their Phase 3 visit date will be assigned a PM_2.5_ exposure calculated using the daily concentrations estimated for the Phase 3 address. If that participant has both Phase 2 and 3 addresses and a between-survey interval starting between those dates, the average of the daily concentrations at the two geocoded address will be used instead. If the participant has a Phase 1 address and the between-survey interval starts between the Phase 1 visit date and 3 years prior to it, the exposure will be calculated using the daily PM_2.5_ concentrations estimated for the Phase 1 address. Other situations follow analogously.

Temperature

Daily minimum and maximum temperatures at 1km resolution were extracted from DayMET at each participants’ Phase 1, Phase 2 and Phase 3 residences ^26^. They were averaged over between-survey intervals using the same method as for PM_2.5_.

Elevation

Elevation at geocoded residences was determined using the elevatr package^27^ in R^28^ version 4.4.2^29^, which uses digital elevation maps from Amazon Web Services Terrain Tiles. Elevation was assigned to between-survey intervals using the same method as for PM_2.5_.

Neighborhood Characteristics

Geocoded locations were used to identify US census tracts in which participants lived and thereby derive their tracts’ 2018 area deprivation index (ADI)^21^, 2018 social vulnerability index (SVI)^22^, and neighborhood’s Homeowners Loan Corporation (HOLC) grade from the 1930s (i.e., redlining grade)^23^. ADI and SVI during each between-survey interval were calculated from the values determined at Phase visits using a similar averaging method to PM_2.5_, while HOLC grade was derived based on the assumed address on the first day of each interval. ADI and SVI were treated as percentiles, while HOLC grade takes the values A, B, C, D, or None, with D being the ‘least desirable’, as determined by property appraisers of the 1930s.

Statistical Analyses

Descriptive Analyses

Statistical summaries were calculated for participants’ demographic profiles, overall health, smoking status, and comorbidities at enrollment, as well as social and environmental exposures. Histograms visualized the distribution of participants’ exacerbation rates and length of between-survey intervals. Following geocoding, all subsequent analyses were performed in R version 4.4.2^28^.

Effect Modification Analyses

To determine whether associations identified in our main model might be impacted by meteorology, time period, neighborhood and demographic factors, and medical history at baseline, we explored the modification of the PM_2.5_ effect. Details These included: 16-year average PM_2.5_, minimum daily temperature, elevation, SVI, ADI, HOLC grade, smoking status, race, sex, age, dichotomous study period (through 2015 or 2016 and later), trichotomous study period (through 2014, 2015-2018, or 2019 and later), as well as FEV_1_/height, % emphysema, pack years of smoking, GOLD stage, asthma diagnosis, prior exacerbations, and Body Mass Index (BMI) at enrollment. To investigate the combined impact of smoking history and disease severity at enrollment, two interaction models with GOLD stage were run, one with GOLD 0 split between current and former smokers, and the other with a single GOLD 0 category. Interaction models included both linear terms and multiplicative interactions with PM_2.5_. Likelihood ratio tests were performed to evaluate model fit with respect to our main model.

Sensitivity Analyses

First, to compare with the method outlined above, PM_2.5_ exposure associated with survey intervals falling between study visits were assigned PM_2.5_ based on the geocoded address at the most recent study visit instead of the average of the most recent and next study visits. Second, since elevation alone may not fully account for oxygen deficit, supplemental oxygen use since the previous survey was included as a covariate. Third, because nonlinear control for time may be overly complex, nonlinear control for time was replaced a simple linear trend, by trichotomous and dichotomous year categories, and by no control for time. Fourth, daily minimum and maximum temperature were (separately) included as covariates due to their potential impact on COPD symptoms. Fifth, because the inclusion of never-smokers in our study population may skew the results, we removed them from our modeling. Sixth, since our results may be skewed by surveys that include unrealistically high exacerbation counts given their between-survey intervals, we remove surveys with an implied rate of ≥20 exacerbations per year. Seventh, to investigate the impact of PM2.5 exposure on the presence of any exacerbations, we replaced the quasi-Poisson model with a quasi-binomial using the presence/absence of an exacerbation in a given between-survey interval as the outcome. Finally, to evaluate the impact of constraining the fixed individual intercepts, the statistical model implemented using gnm was replaced by a random intercept model implemented in the lme4 package^33^. Additionally, since participants may not accurately recall the number of exacerbations they experienced since the previous phone survey, especially over longer intervals, the analysis was repeated using survey intervals less than 18 months, 15 months, 12 months, and 9 months.

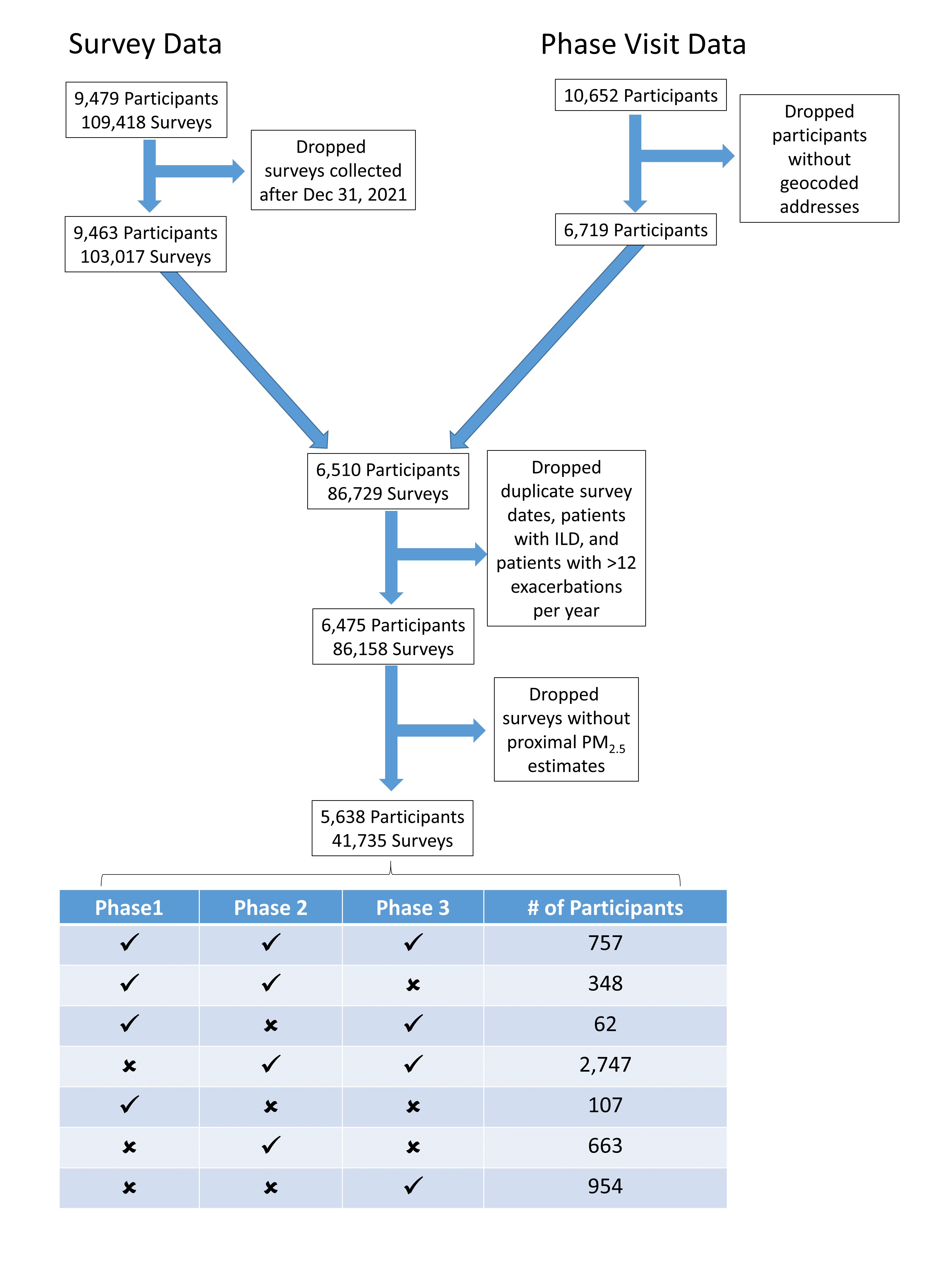

Supplemental Figure S1: Data processing flowchart

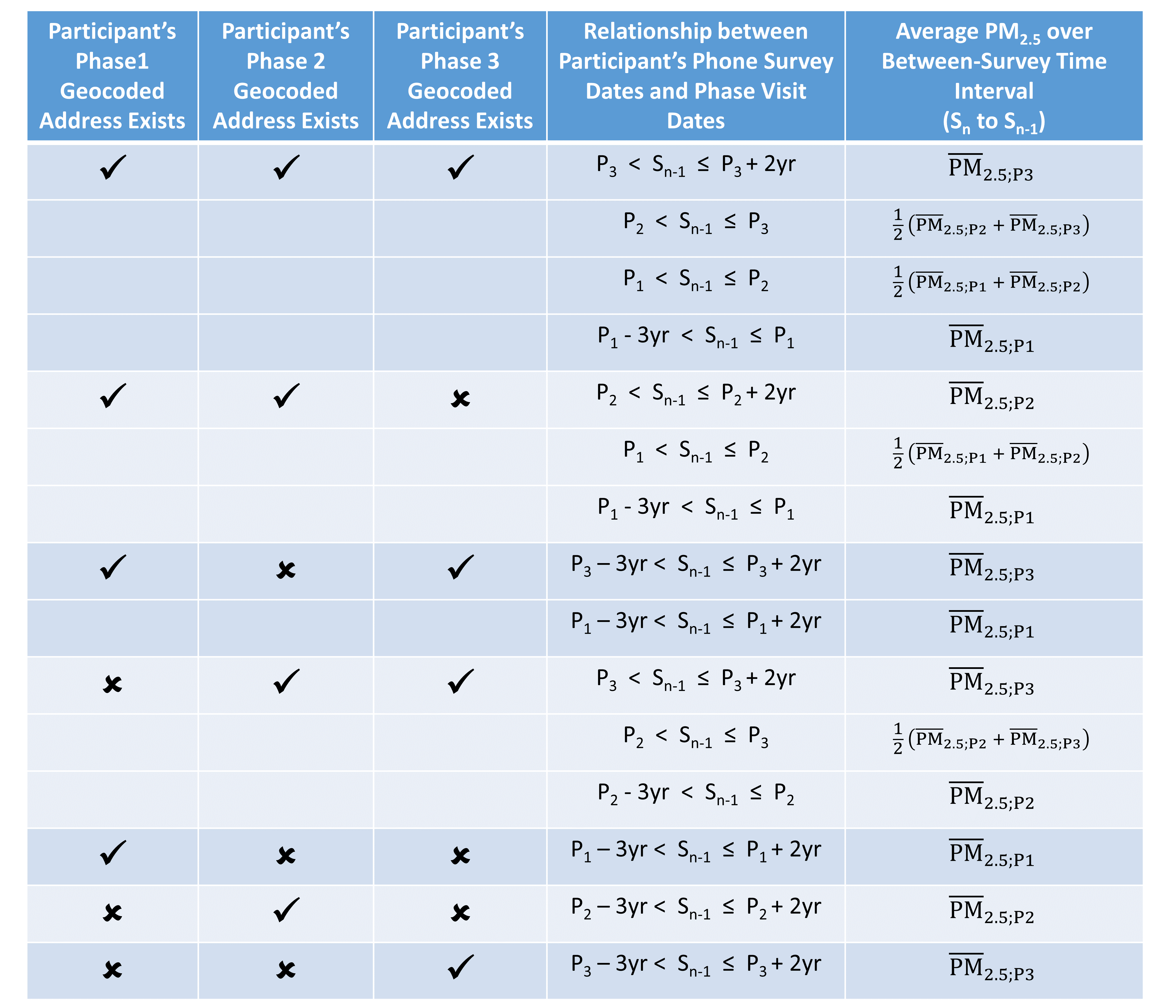

Supplemental Table 1: Exposure assignment table used to link daily PM2.5 levels interpolated to geocoded addresses at phase visits to dates of phone surveys in order to calculate the participant’s average PM2.5 during each between-survey interval. P1, P2, and P3 represent the phase visit dates for a given participant. S_n_ and S_n-1_ represent the date of the n-th phone survey date and of the previous survey, respectively (enrollment is considered S_0_). The table is consulted independently for each survey interval for each participant.

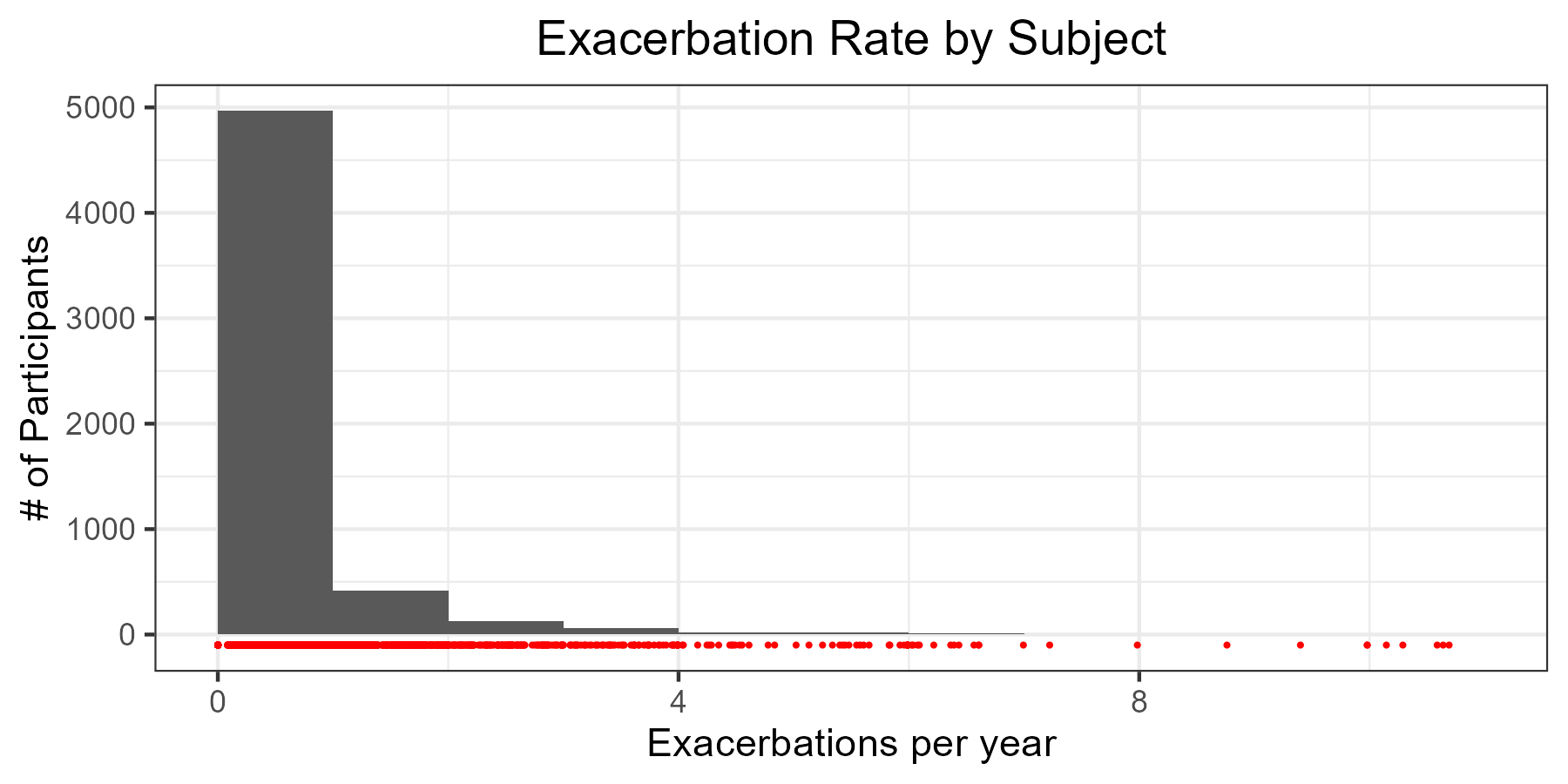

Supplemental Figure S2: Exacerbation rate histogram

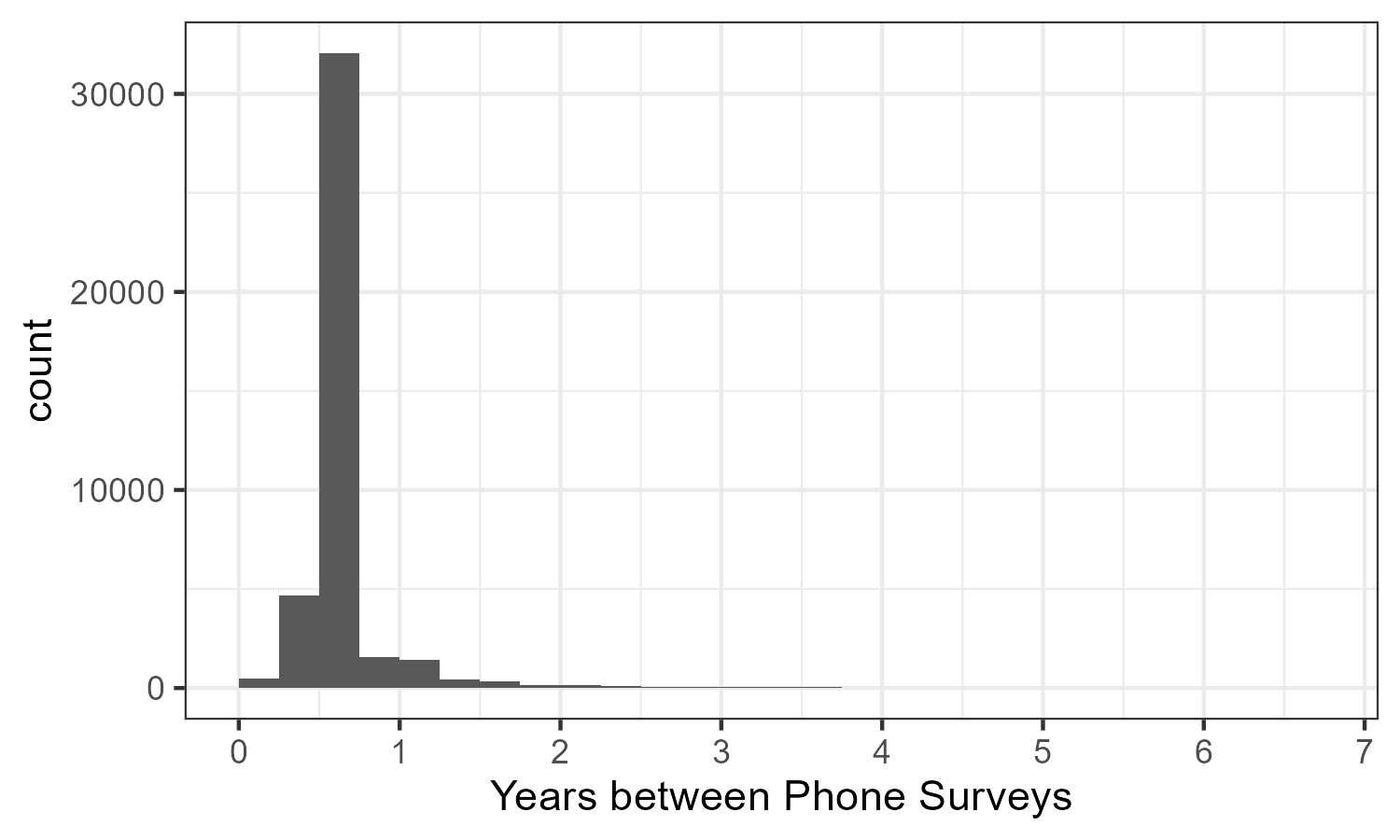

Supplemental Figure S3: Time intervals between surveys and most recent patient contacts (surveys or study visits)

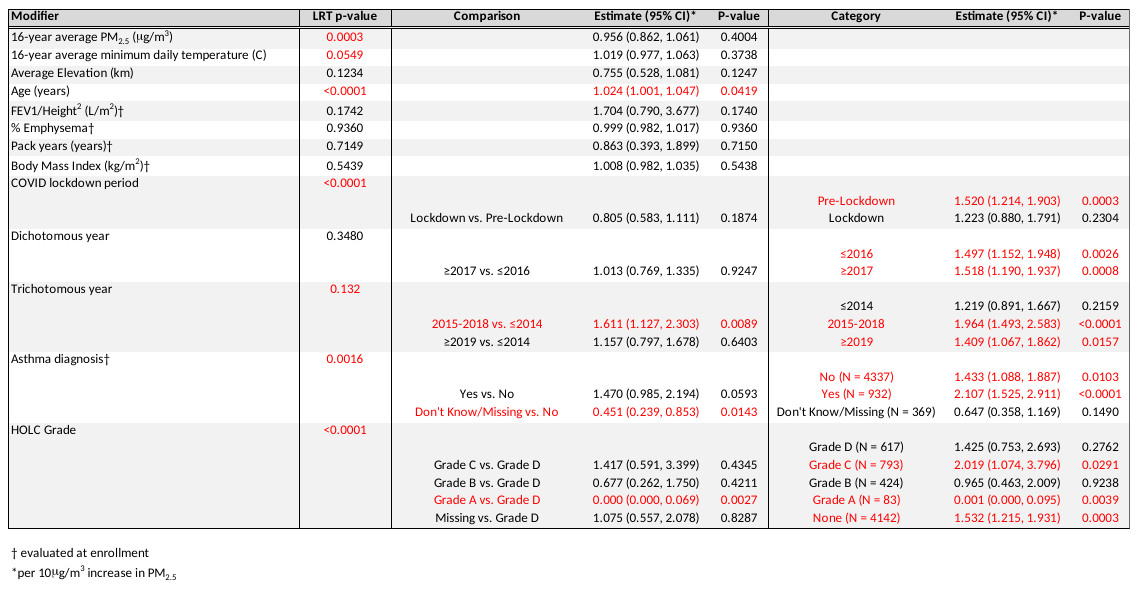

Supplemental Table S2: Effect modification results. Results with p<0.05 are shown in red.

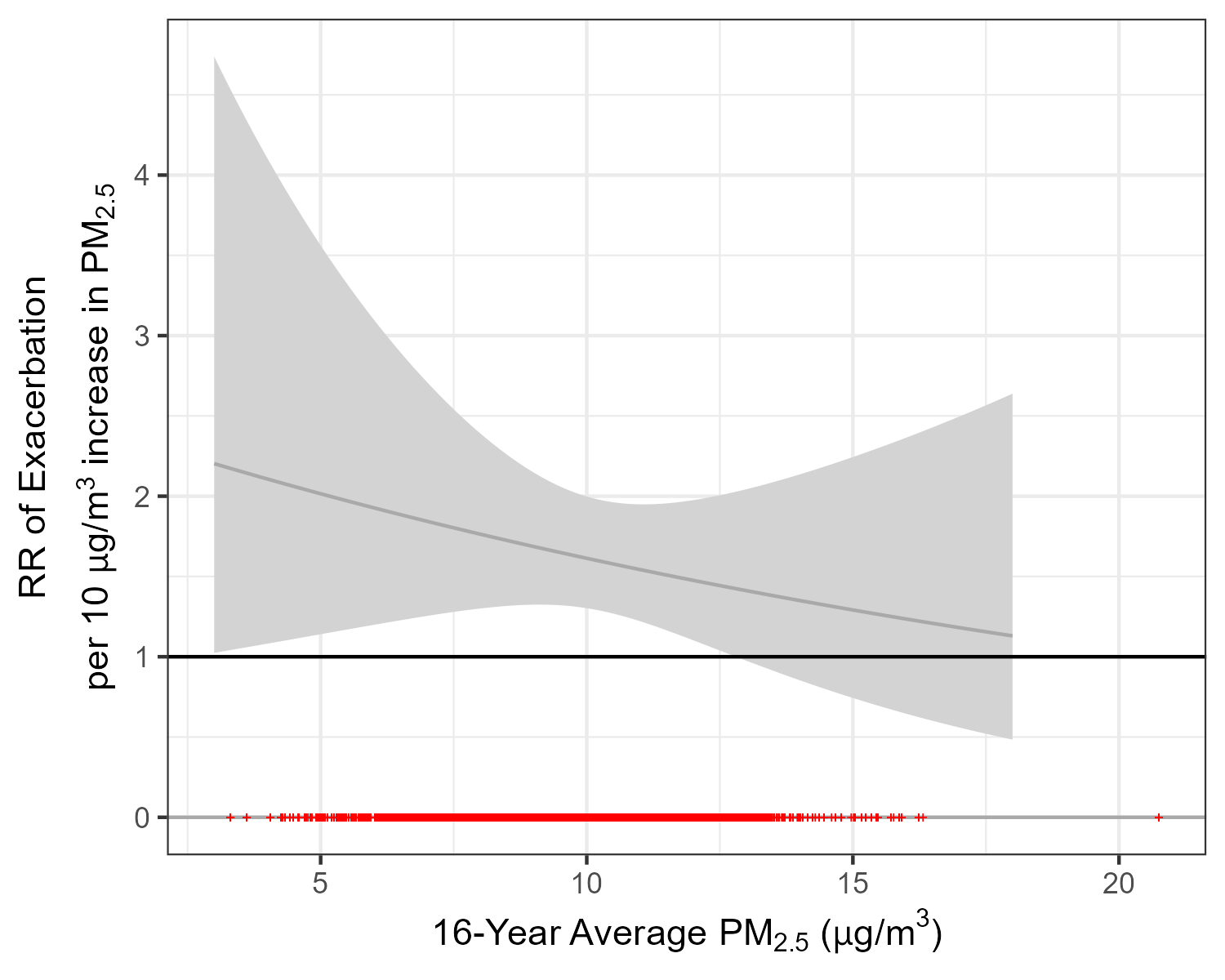

Supplemental Figure 4: Variation in the PM_2.5_-excerbation association by 16-year average daily PM_2.5_ concentration at residence

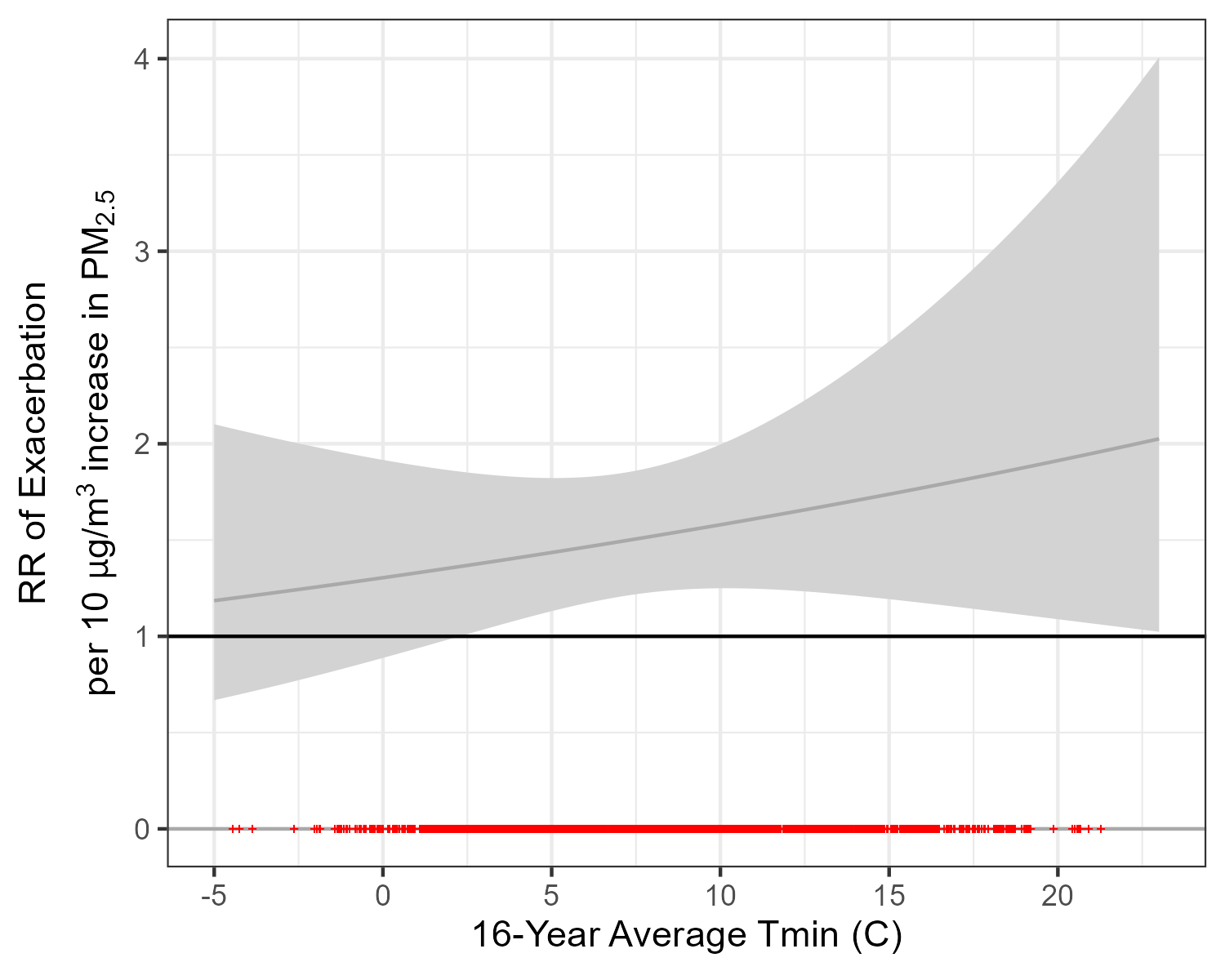

Supplemental Figure 5: Variation in the PM_2.5_-excerbation association by 16-year average daily minimum temperature at residence

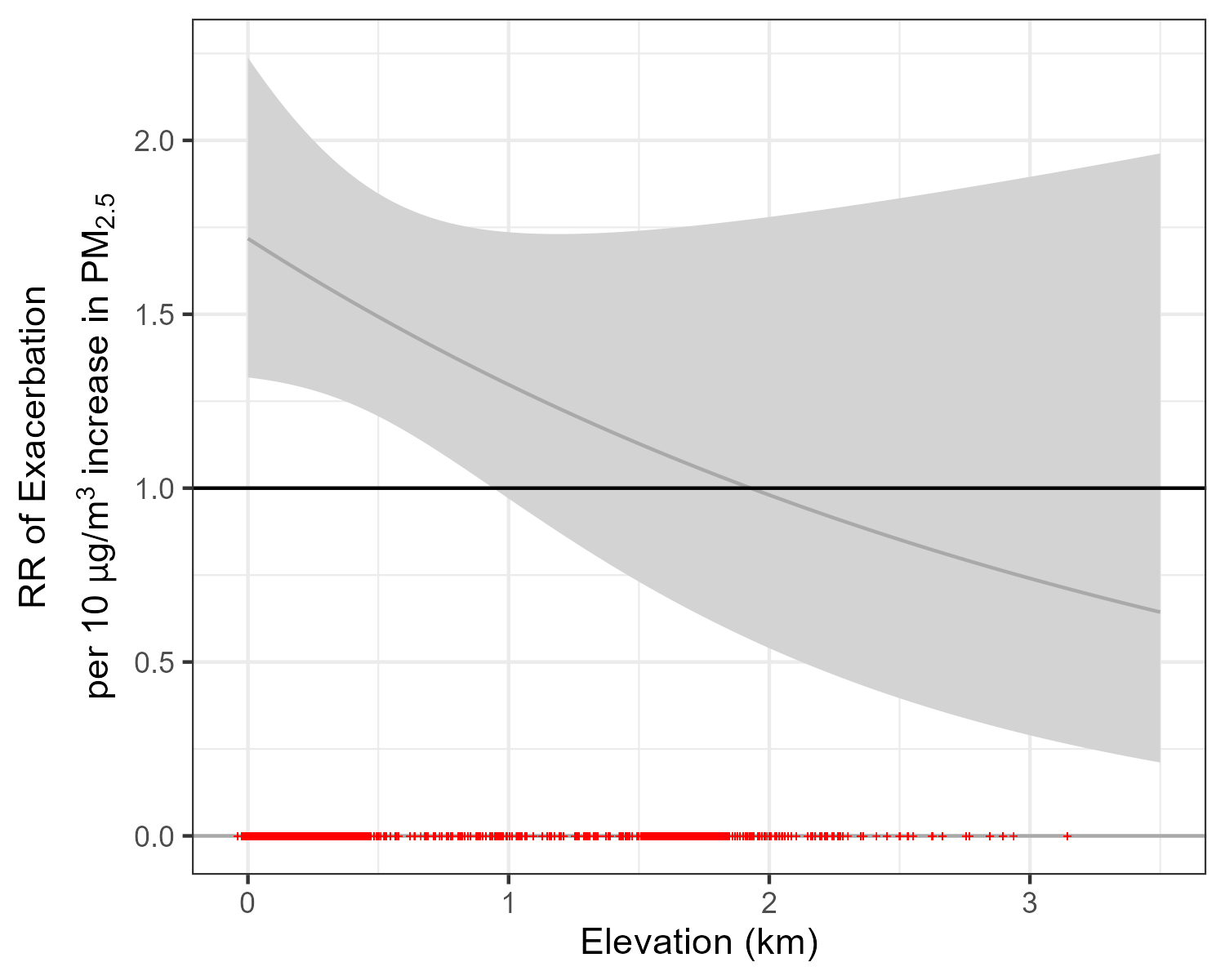

Supplemental Figure 6: Variation in the PM_2.5_-excerbation association by elevation

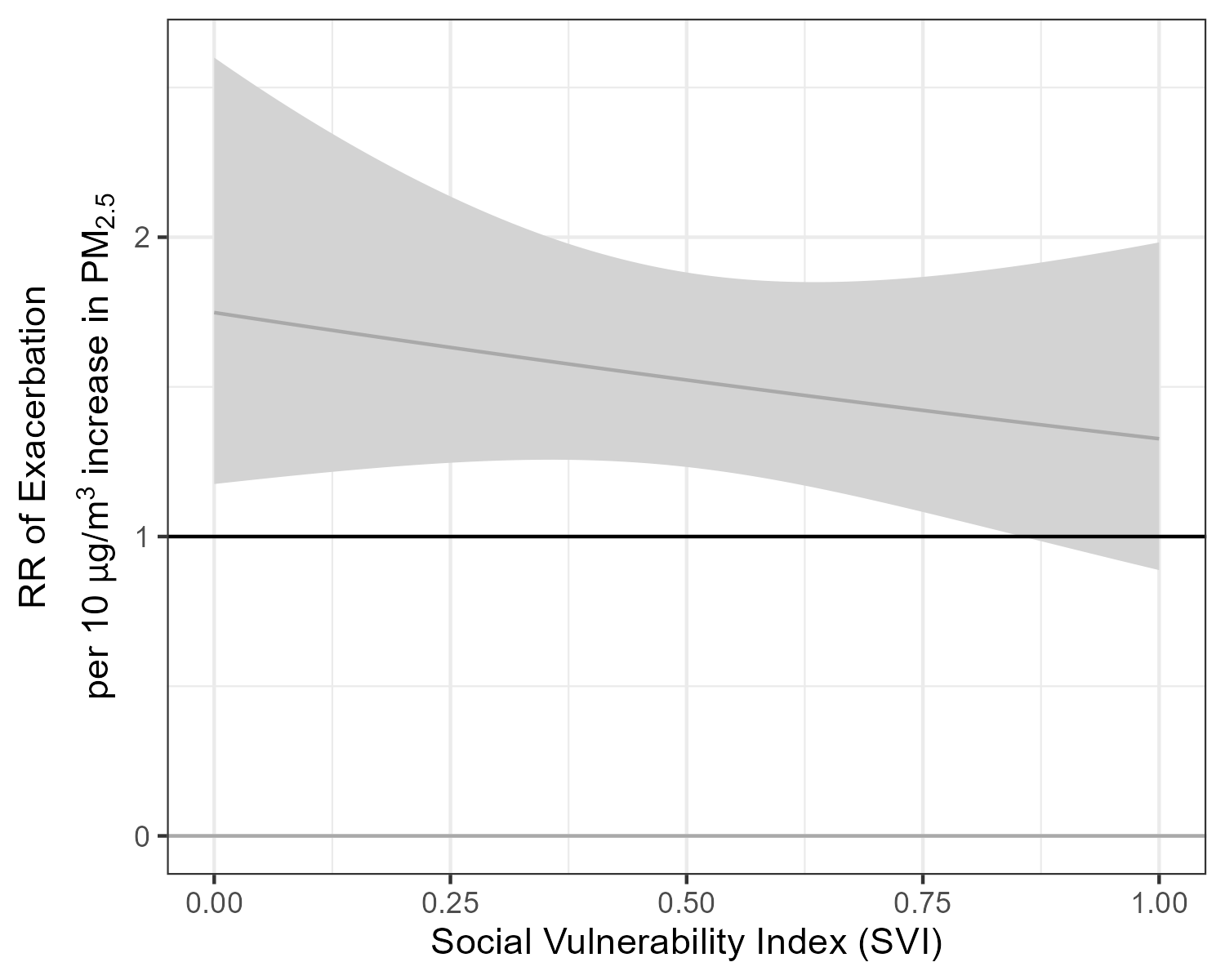

Supplemental Figure 7: Variation in the PM_2.5_-excerbation association by Social Vulnerability Index

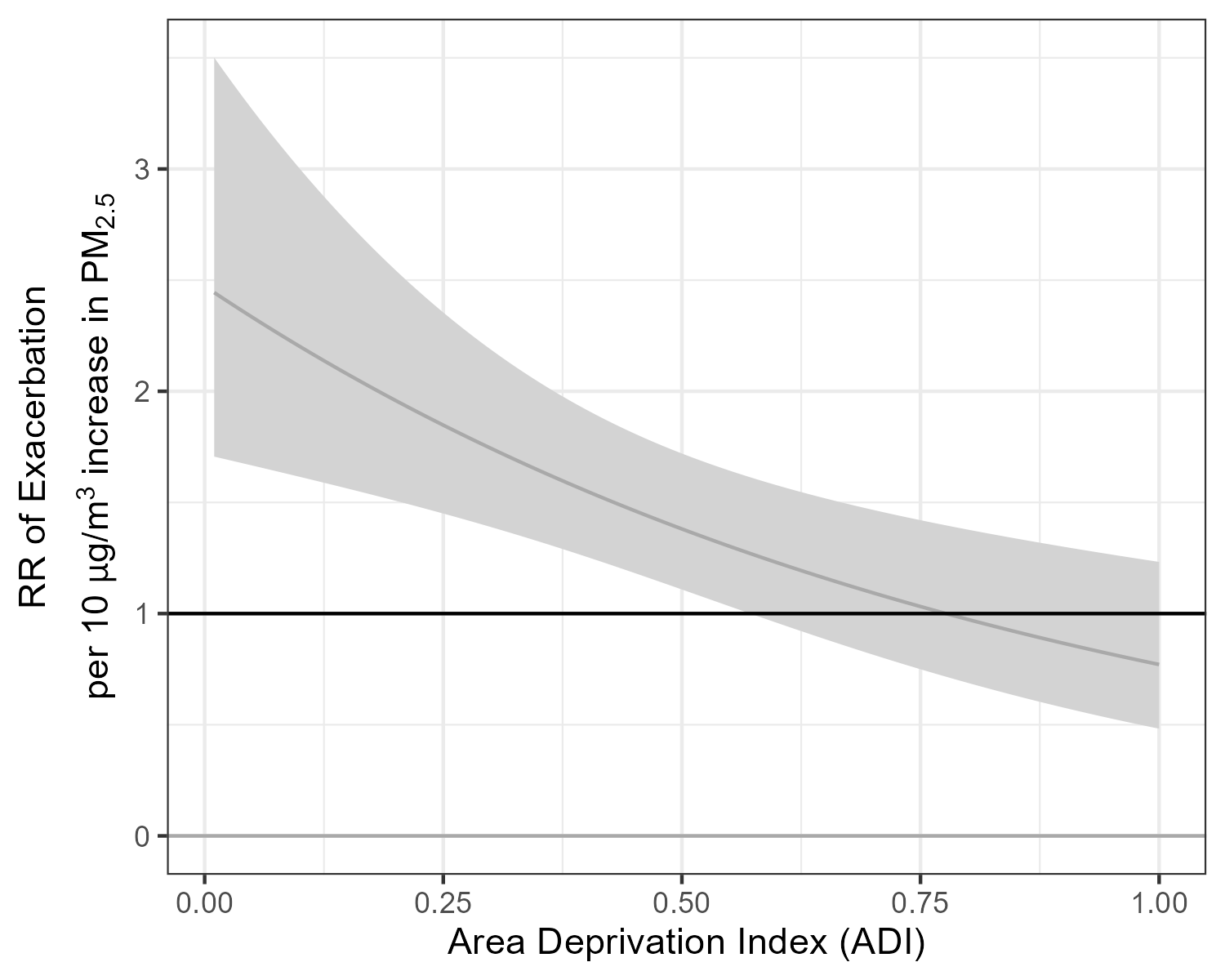

Supplemental Figure 8: Variation in the PM_2.5_-excerbation association by Area Deprivation Index

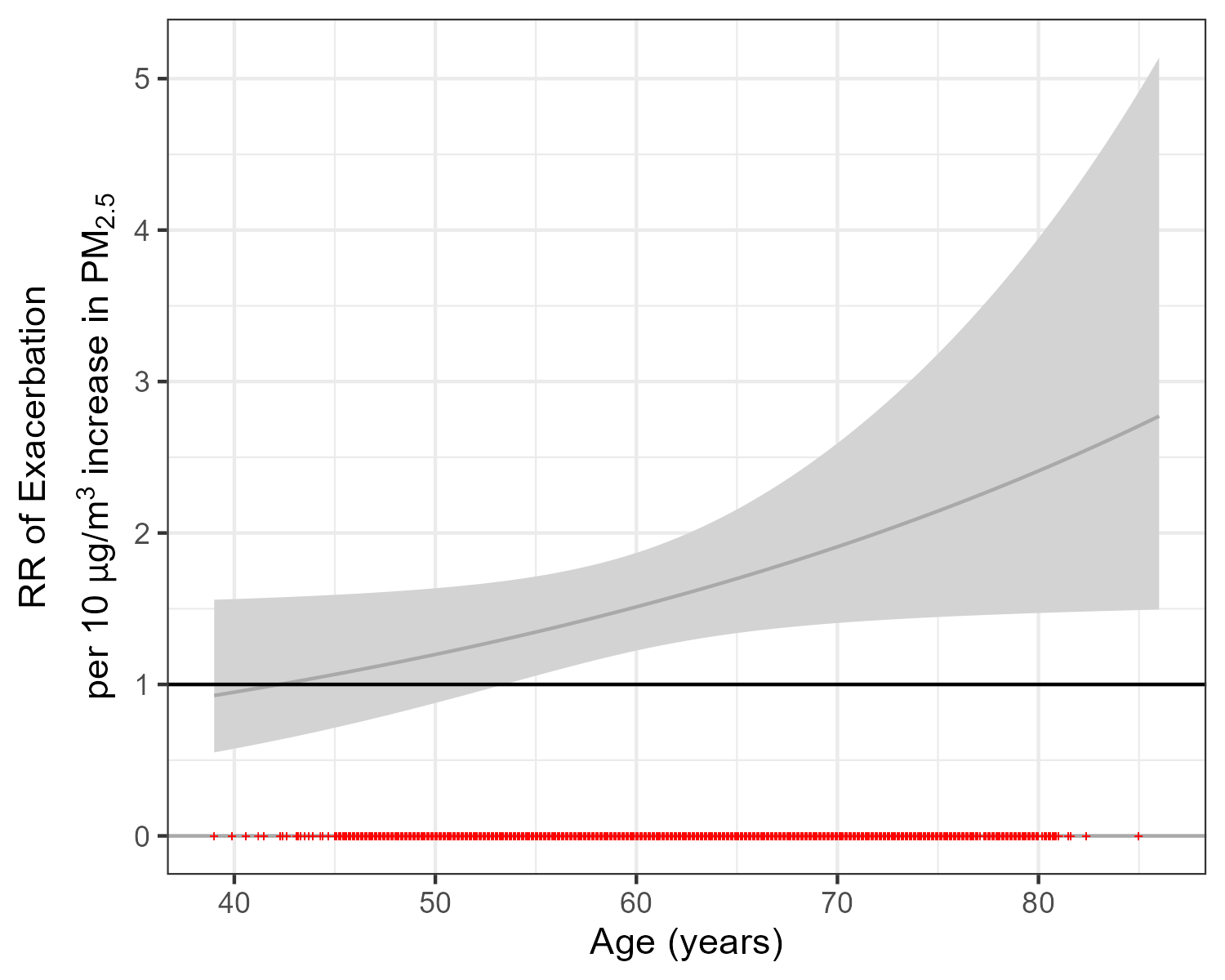

Supplemental Figure 9: Variation in the PM_2.5_-excerbation association by age

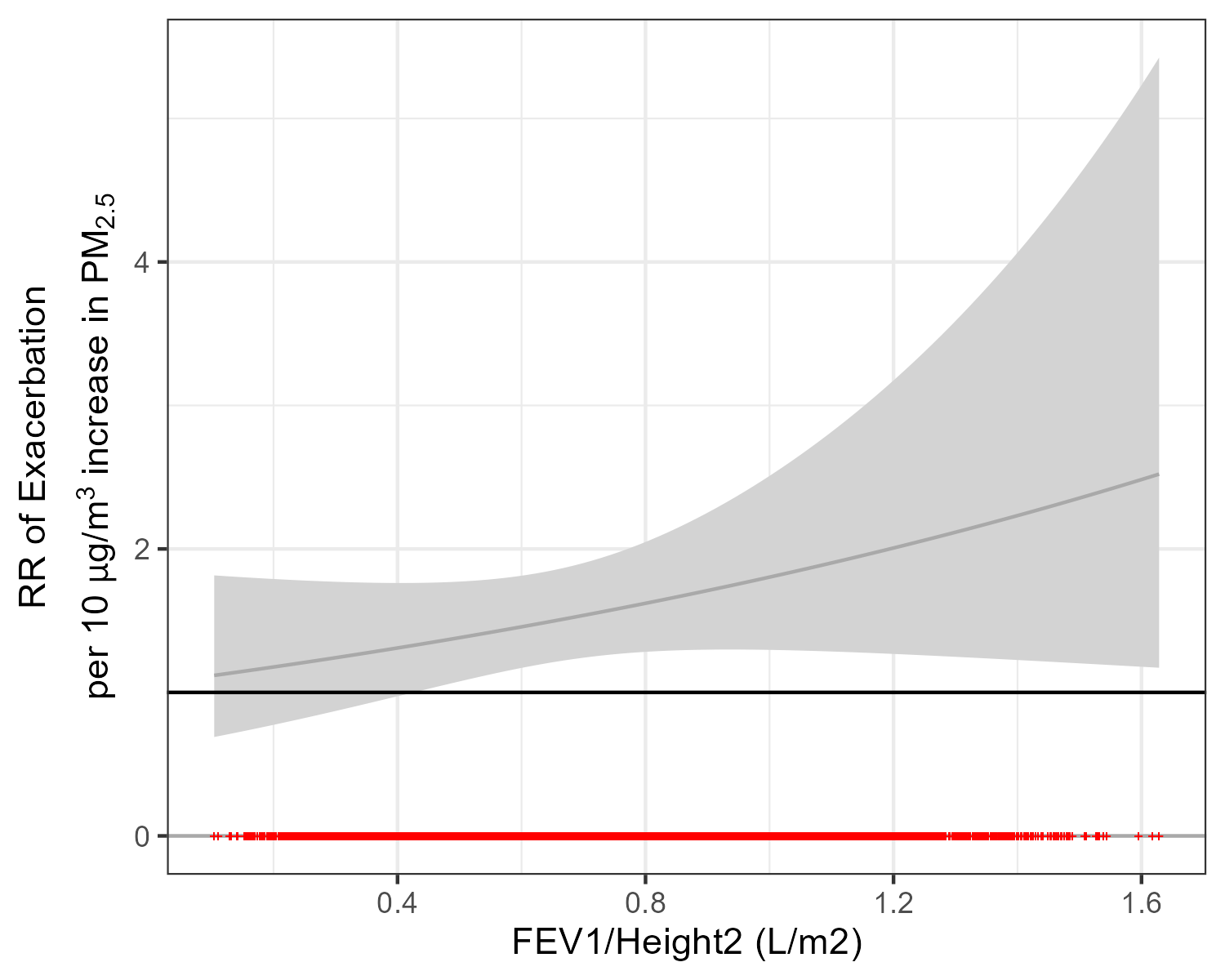

Supplemental Figure 10: Variation in the PM_2.5_-excerbation association by FEV1/height

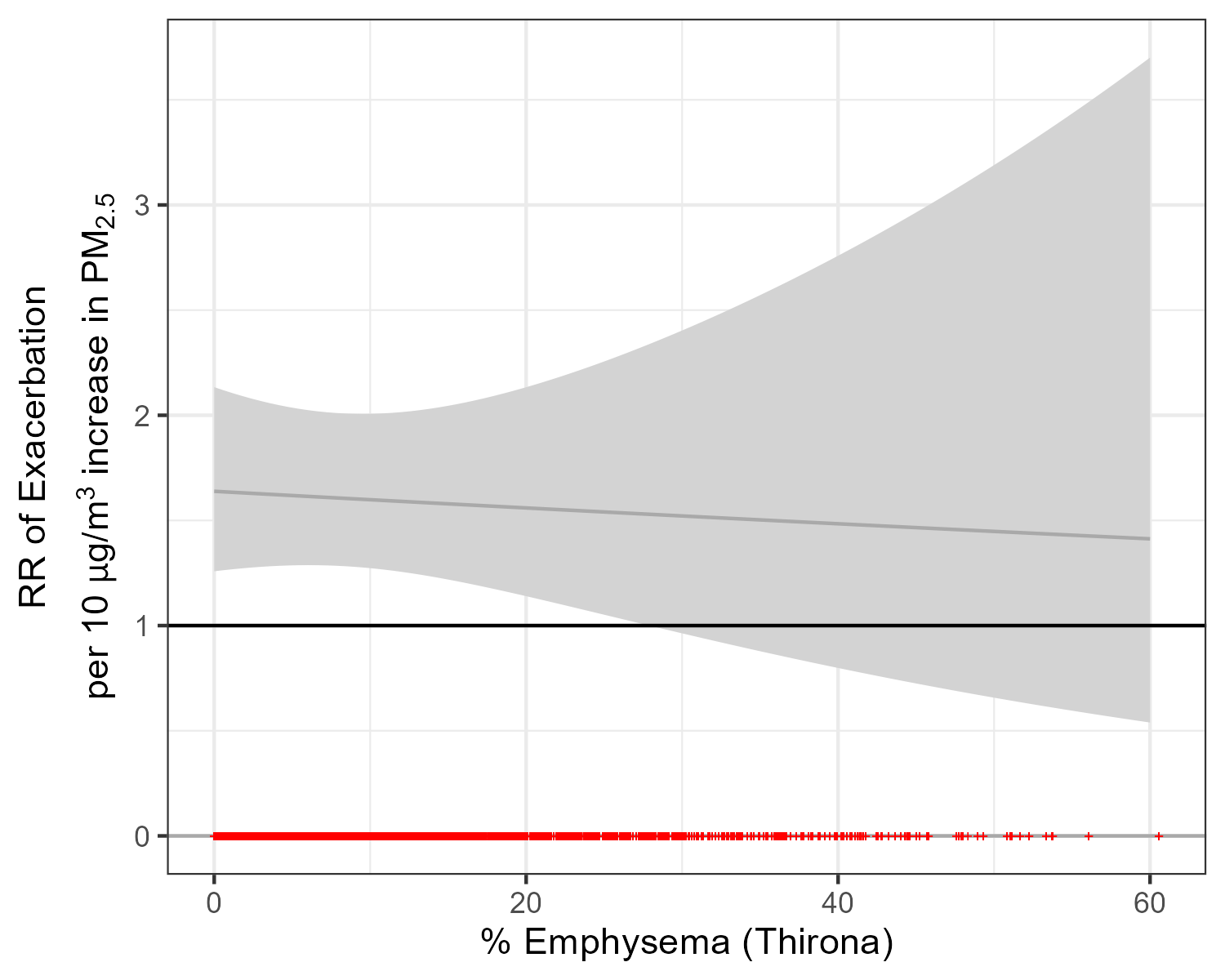

Supplemental Figure 11: Variation in the PM_2.5_-excerbation association by % emphysema

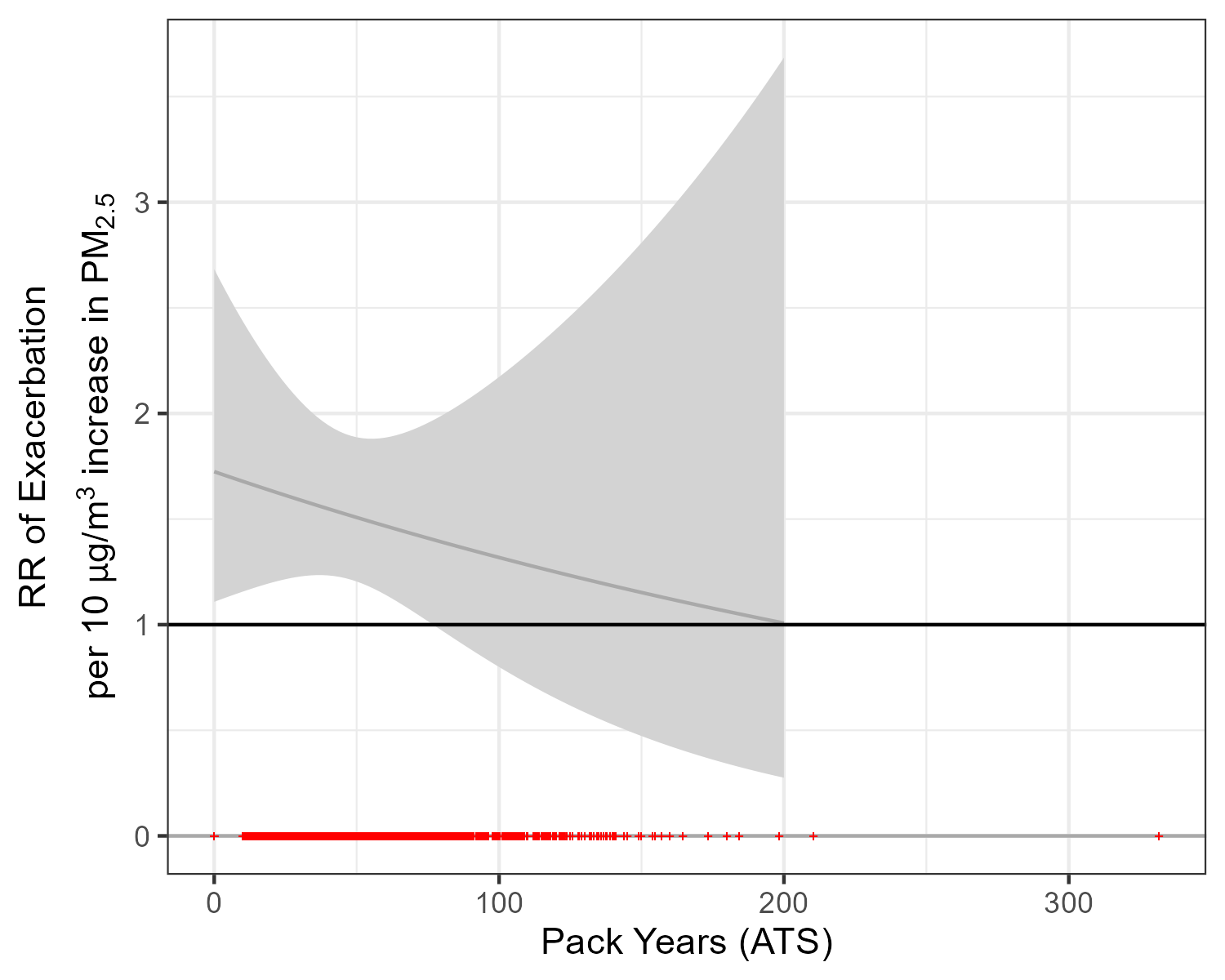

Supplemental Figure 12: Variation in the PM2.5-excerbation association by pack years smoked

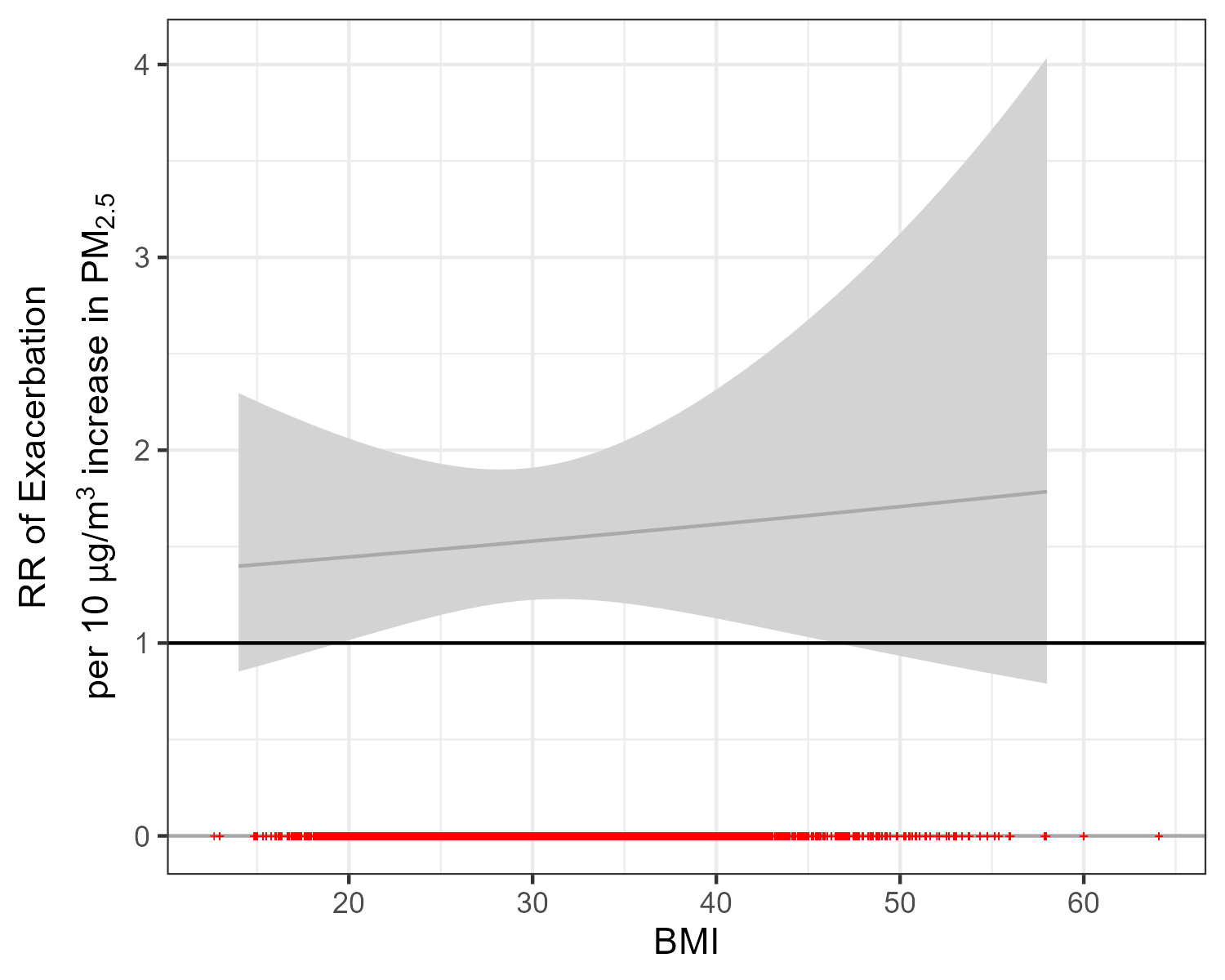

Supplemental Figure 13: Variation in the PM2.5-excerbation association by Body Mass Index

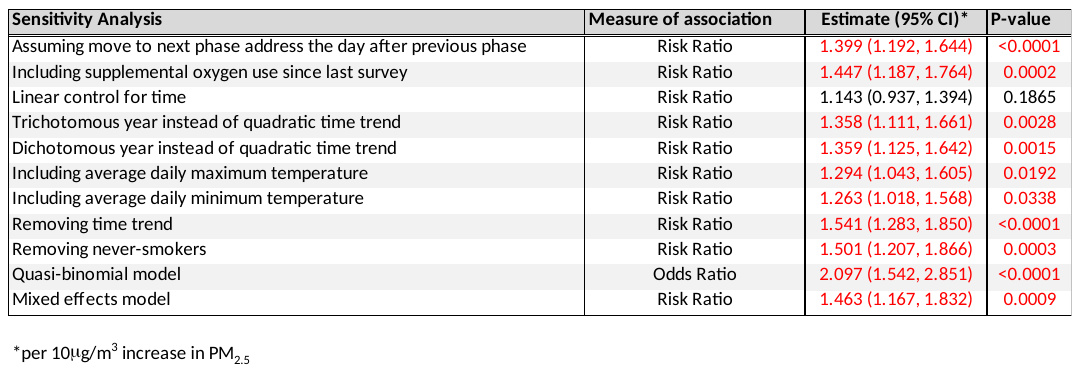

Supplemental Table S3: Sensitivity analysis results. Results with p<0.05 are shown in red.

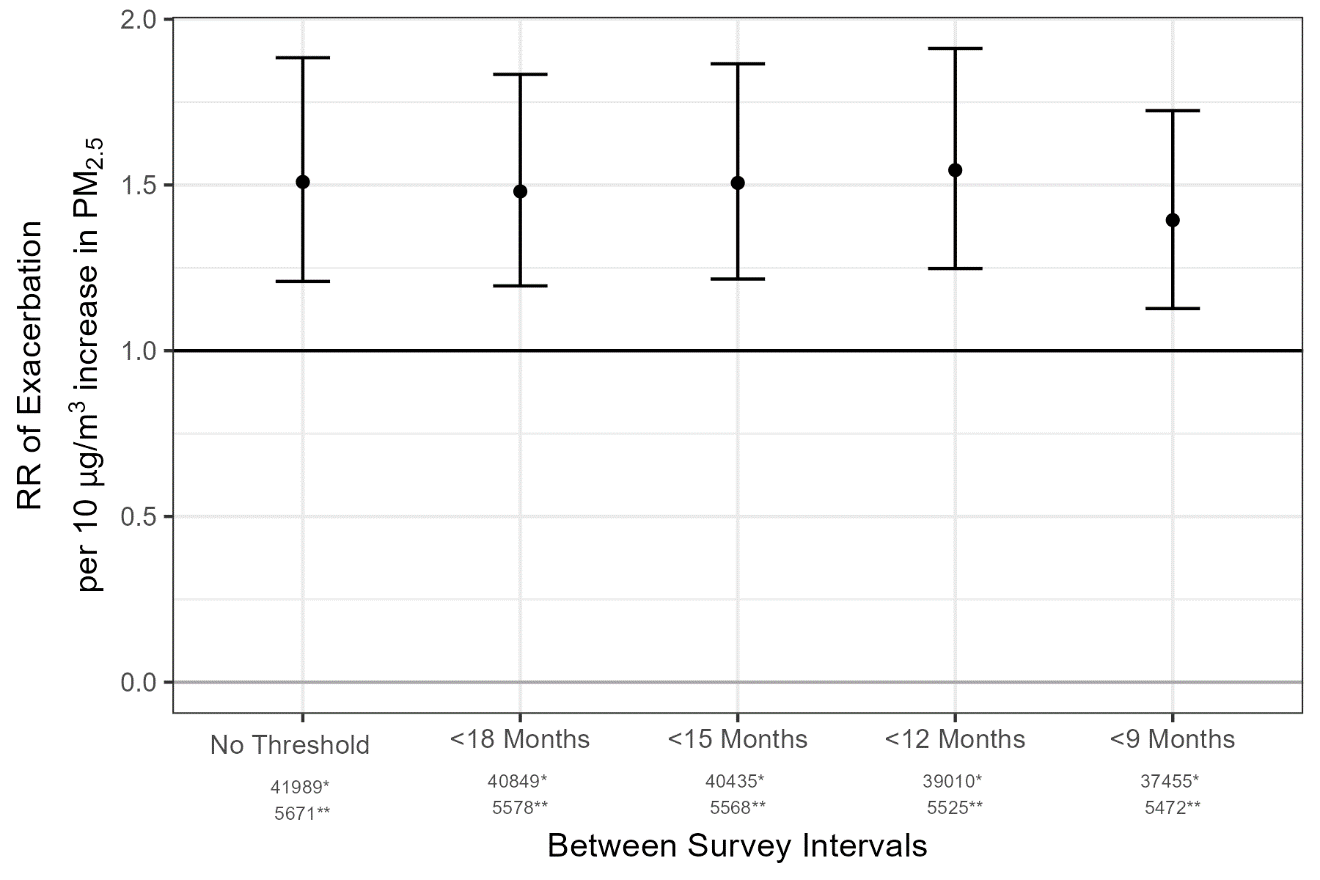

Supplemental Figure S14: Results thresholded by the time interval between patient contacts.

* = number of surveys after thresholding

** = number of participants after thresholding.

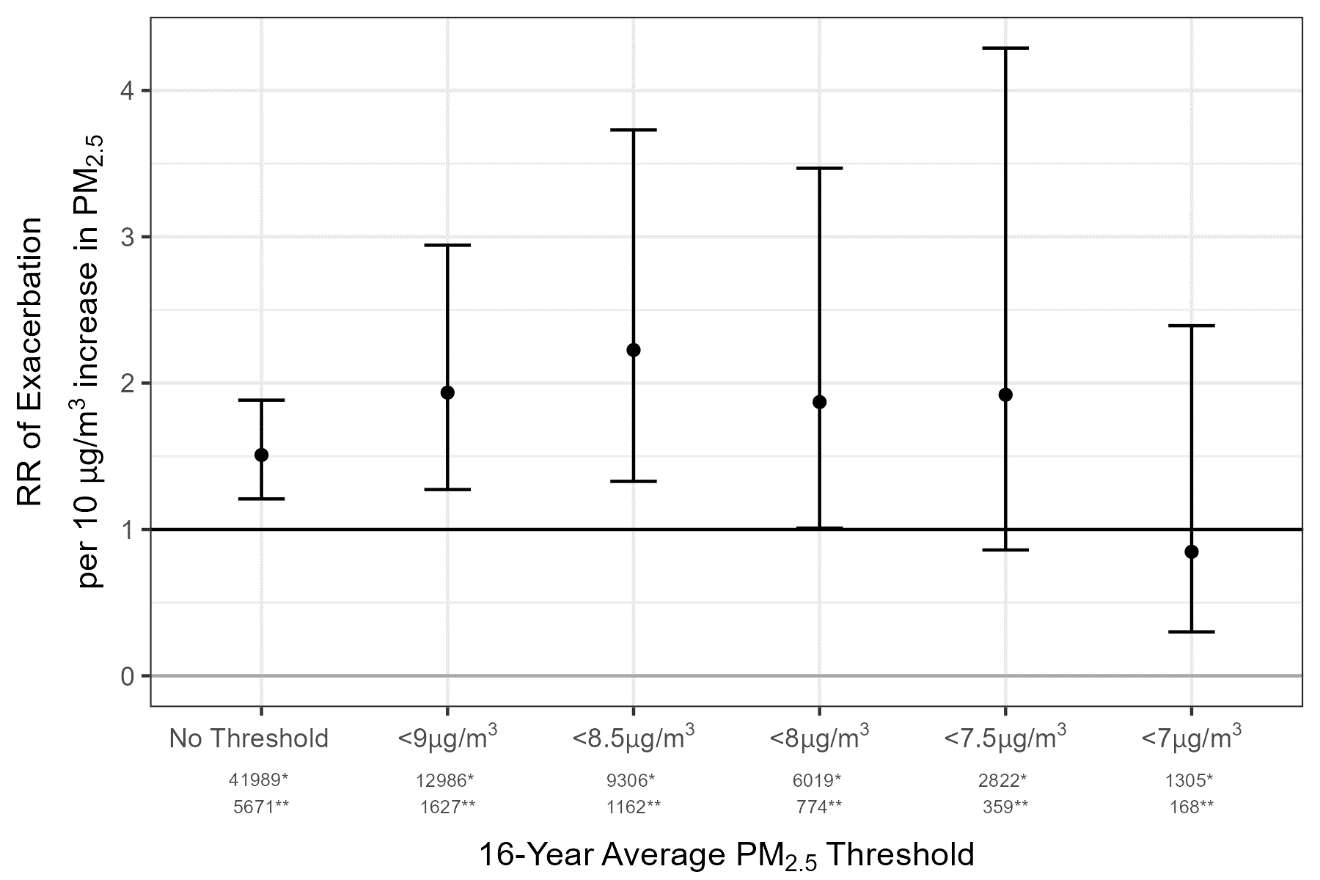

Supplemental Figure S15: Results thresholded by long-term average PM_2.5_ concentration as residence.

* = number of surveys after thresholding

** = number of participants after thresholding.

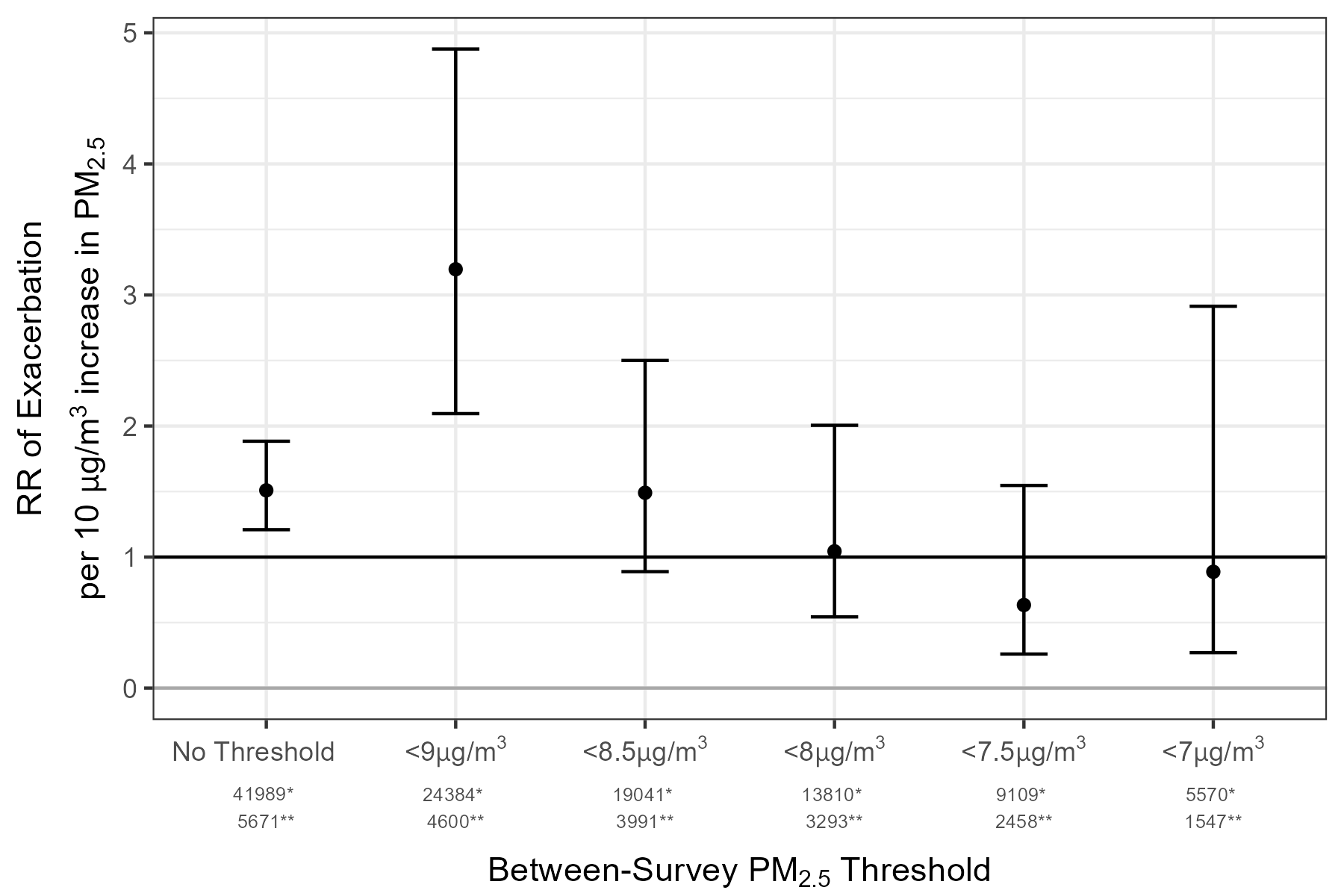

Supplemental Figure S16: Results thresholded by long-term average PM_2.5_ concentration as residence.

* = number of surveys after thresholding

** = number of participants after thresholding.
